# Genetic scores explain variation in birthweight that is not captured by easily measured clinical and anthropometric variables

**DOI:** 10.1101/2022.02.10.22270369

**Authors:** Maneka Haulder, Alice E Hughes, Robin N Beaumont, Bridget A. Knight, Andrew T. Hattersley, Beverley M Shields, Rachel M Freathy

## Abstract

**Background:** Human birthweight is a complex, multifactorial trait. Maternal characteristics contribute to birthweight variation by influencing the intrauterine environment. Variation explained by genetic effects is also important, but their contributions have not been assessed alongside other key determinants. We aimed to investigate variance in birthweight explained by genetic scores in addition to easily-measurable clinical and anthropometric variables.

**Methods:** We analysed 549 European-ancestry parent-offspring trios. We investigated variance explained in birthweight (adjusted for sex and gestational age) in multivariable linear regression models including genetic scores, routinely-measured maternal characteristics and parental anthropometric variables. We used R-Squared (R^2^) to estimate variance explained, adjusted R-squared (Adj-R^2^) to assess improvement in model fit from added predictors, and F-tests to compare nested models.

**Results:** Maternal and fetal genetic scores together explained 6.0% variance in birthweight. A model containing maternal age, weight, smoking, parity and 28-week fasting glucose explained 21.7% variance. Maternal genetic score explained additional variance when added to maternal characteristics (Adj-R^2^ =0.233 vs Adj-R^2^=0.210, p<0.001). Fetal genetic score improved variance explained (Adj-R^2^=0.264 vs 0.248, p<0.001) when added to maternal characteristics and parental heights.

**Conclusions:** Genetic scores account for variance explained in birthweight in addition to easily measurable clinical variables. Parental heights partially capture fetal genotype and its contribution to birthweight, but genetic scores explain additional variance. While the genetic contribution is modest, it is comparable to that of individual clinical characteristics such as parity, which suggests that genetics could be included in tools aiming to predict risk of high or low birthweights.

**Key messages:**

- Known contributors to variation in birthweight include (i) factors associated with the maternal intrauterine environment (e.g. maternal glycaemia or smoking), and (ii) parental heights, which capture some of the genetic contribution to fetal growth. However, the added contribution of genetic scores composed of common birthweight-associated variants has not been assessed.
- We showed, using 549 parent-offspring trios, that maternal and fetal genetic scores explained additional variation in sex-and gestational age-adjusted birthweight, when added to maternal variables that are easily obtained in a clinical setting (age, weight, smoking, parity and 28-week fasting glucose).
- Parental heights explained variance in birthweight independently of routinely measured maternal clinical variables, but the maternal and fetal (or paternal) genetic scores made additional, independent contributions to birthweight variance.
- The genetic score contribution was modest, but it was comparable to that of individual clinical characteristics such as parity, which suggests that genetics could be included in tools aiming to predict risk of high or low birthweights.
- Since this work was limited to a UK sample of European ancestry, it will, however, be important to test the relative contributions of genetics and other factors to birthweight variation in diverse populations.

## Introduction

Birthweight is a complex trait with considerable variability. It is important to understand what contributes to this variability because babies born large for gestational age (LGA) or small for gestational age (SGA) are at a higher risk for adverse pregnancy and perinatal outcomes^1^. There are also well replicated associations between variation in birthweight and risks of later life cardio-metabolic disease^2, 3^.

Previous research has shown that factors associated with the maternal intrauterine environment, for example, maternal glycaemia, age, parity, weight and smoking, account for some variation in birthweight, once fetal sex and gestational duration have been accounted for ^4^. Maternal smoking during pregnancy is associated with lower birthweight^5^. Parity is also associated with birthweight ^6^, with babies of later birth order having higher birthweight, on average. A low pre-pregnancy BMI increases the risk of SGA and a high pre-pregnancy BMI has been found to increase the risk of LGA ^7^. There is a positive continuous association between maternal fasting glucose and birthweight ^8^. However, each of these variables contributes only modestly to birthweight variation. For example, maternal fasting glucose levels have been reported to explain only a small fraction (10%) of variation in birthweight^9^, and most LGA babies are not born to mothers with glucose levels that are high enough to be classified as diabetes^10^.

Fetal genetic variation contributes to variation in birthweight independently of the intrauterine environment and is therefore important to consider. Some of the fetal genetic contribution to birthweight can be captured by measuring paternal or maternal height. Height is a highly heritable trait, and the correlation between birthweight and paternal height in particular, via fetal skeletal growth ^11^ occurs due to genetic inheritance.

A recent genome-wide-association study (GWAS) identified 190 regions of the genome where common single nucleotide polymorphisms (SNPs) are associated with birthweight variation^12^. The associated genetic variants at three-quarters of the 190 identified loci exert their effects directly from the fetal genotype, with a small proportion of those showing additional maternal effects. Associated variants at the other quarter of identified loci originated from the mother’s genome and showed indirect effects, via the maternal environment. A fetal genetic score consisting of 58 variants was shown to make a significant contribution to birthweight independently of maternal glucose levels^13^, suggesting measurements of fetal genetics could add to the variance in birthweight explained by other factors. However, the contribution of genetic variation to birthweight has not been assessed directly alongside other clinical variables. We therefore aimed to assess the contributions of genetic scores to variation in offspring birthweight, in addition to easily obtained clinical and anthropometric variables, in a UK community-based study of mothers, fathers and children.

## Methods

### Study population

We used data from the Exeter Family Study of Childhood Health (EFSOCH)^14^. EFSOCH is a study based on children born between 2000 and 2004 in postcodes EX1-4 in central Exeter, UK.

Inclusion criteria for the current analyses consisted of only those parent-offspring trios where the offspring was born at term (≥37 and <42 weeks gestation^14^) and complete clinical, anthropometric and maternal, paternal and fetal genetic data were available. Most trios had complete phenotype data, but following genotype quality control, and owing mainly to lower availability of fetal DNA from cord blood compared with parental DNA, complete genotype data was available for both parents and offspring in 60% of the trios. The final dataset consisted of 549 parent-offspring trios. The selection of variables is illustrated by the flowchart in Figure 1. To check for any differences between excluded and included participants, we used t-tests to compare means of continuous variables of the excluded with the included (maternal height, maternal weight, gestational duration, birthweight and maternal age), and chi-square tests to compare the excluded categorical variables with the included categorical variables (maternal smoking status, parity and sex of the baby).

**Fig 1:**
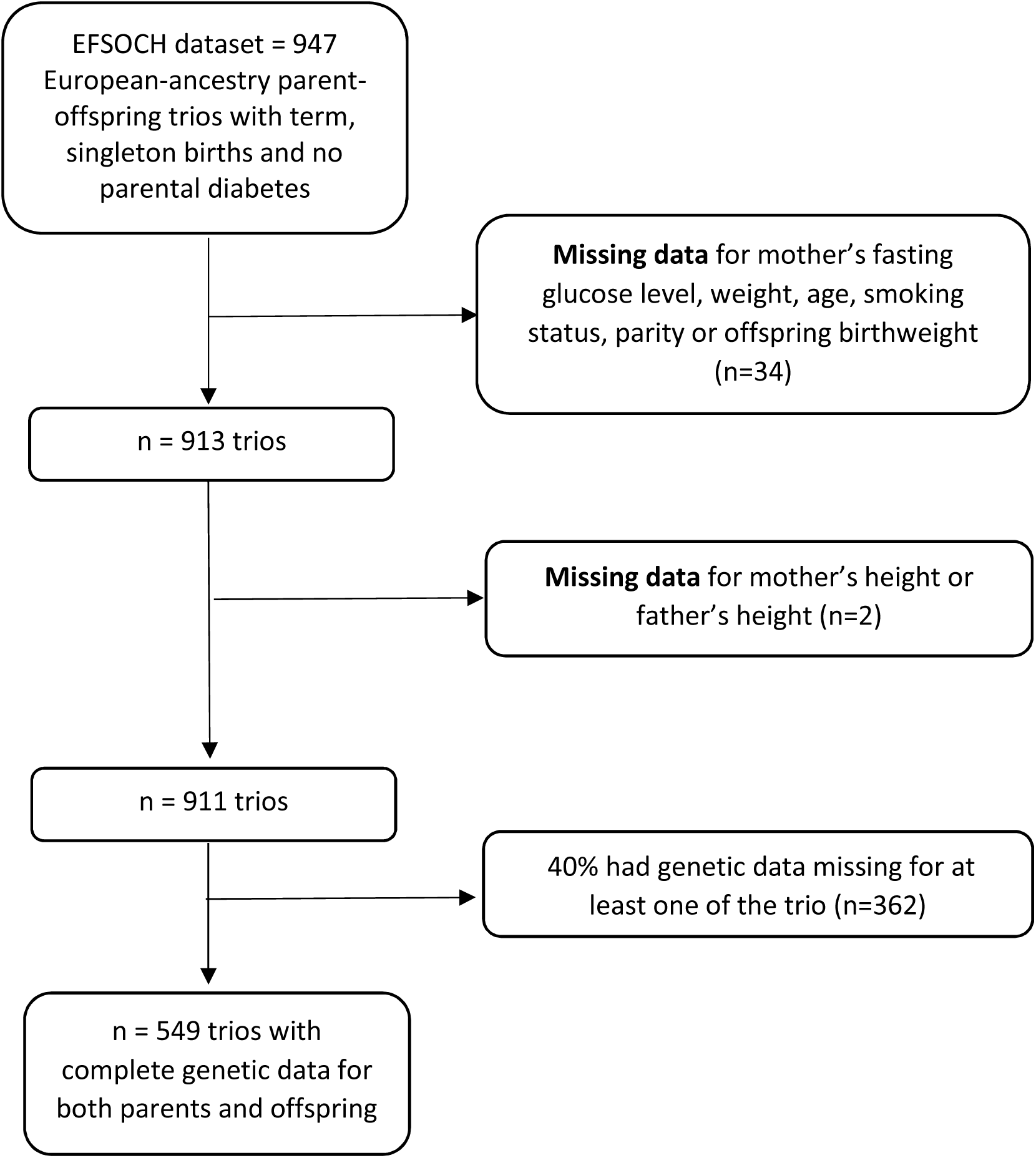
Flowchart illustrating how the data was prepared for analysis

### Characteristics of participants

Full details of data collection are found in the EFSOCH study protocol^14^. Briefly, detailed anthropometric measurements and biochemistry from the parents were taken at 28 weeks’ gestation. All measurements were taken three times and an average value was calculated. Maternal and paternal heights were measured to the nearest 0.1 cm. Maternal weight was measured to the nearest 100 g. Birthweights of the parents were self-reported. Offspring birthweight was measured at birth, to the nearest 10 g and adjusted for sex and gestational age, centred around 40 weeks, according to the UK 1990 birthweight standards ^15^. Maternal glucose was measured in fasting maternal samples (fasting for at least 10h prior to sampling), early morning at the parents’ home. Pregnancy details such as parity were obtained from medical records. Information about the mother’s smoking status was obtained via a questionnaire completed by the mother at recruitment.

### Genotyping

Parental and offspring DNA were extracted to allow molecular genetic analysis of variants implicated in fetal growth. At birth, a sample of cord blood was taken at delivery. DNA was extracted from the spun white cells. The EFSOCH sample (consisting of 2,768 samples: mothers (n=969), fathers (n=937) and offspring (n=862)) genotyping was carried out using the Illumina HumanCoreExome array. A total of 106 samples were excluded due to low call rate, kinship errors, sex mismatches or ancestry outliers. The 2662 included samples were of European ancestry (assessed using flashPCA ^16^ with genotype call rates >98% and phenotypic sex and kinship were validated using genotype data assessed by KING software^17^). The included genotyped SNPs had call rates >95%, Hardy-Weinberg p > 1 × 10^−6^, and minor allele frequency (MAF) >1%. The Haplotype Reference Consortium (HRC) version r1.1 reference panel (Michigan Imputation Server) was used to impute the genotypes in all samples. A total of 98% of the SNPs included in the scores had an imputation quality > 0.4 and a Minor Allele Frequency > 0.001 in EFSOCH (see supplementary **Table S1**).

## Statistical Analyses

### Genetic scores

We created independent maternal and fetal genetic scores for birthweight, and also a paternal genetic score for father’s own birthweight (analogous to a fetal genetic score). We calculated the genetic scores (GS) according to Equation 1, where NSNP is the total number of SNPs, wi is the weight for SNP i and gi is the genotype dosage at SNP i.

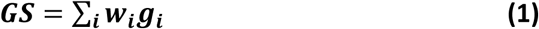

A total of 209 SNPs, identified at 190 loci in the most recent GWAS of birthweight^12^, were used to calculate the maternal, paternal and fetal genetic scores (see **Table S1**). Effect estimates for each SNP were used as weights, and for the maternal score, these had been adjusted to represent the maternal effects independent of fetal genotype effects using a structural equation model^12^. For the fetal score, fetal effect estimates independent of maternal genotype effects were used as weights, and for the paternal score for father’s own birthweight, the fetal GWAS weights were unadjusted so as to capture maximum information. Each genetic score variable was then standardized to a mean of 0 and SD of 1. To validate the genetic scores, we tested the associations between each standardized genetic score and its respective phenotype using simple linear regression models.

### Linear regression models to estimate variance in adjusted birthweight by genetic and other factors

We used multivariable linear regression models to model the variance in birthweight explained by several clinical, anthropometric and genetic factors. We ensured that the regression model assumptions were met and the model assumptions were checked using diagnostic plots of residuals and fitted values. To determine the additional variability explained by genetics, we examined the following models, with birthweight (adjusted for sex and gestational age) as the outcome variable:

**Model 1: Genetic scores model:** maternal and fetal genetic scores were included as predictors to investigate their contribution to birthweight.

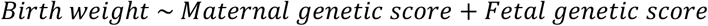

**Model 2: Maternal clinical features (intrauterine environment) model:** maternal fasting glucose, age, weight, parity and the mother’s smoking status were used in this model.

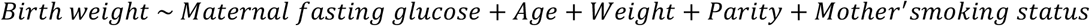

**Model 3: Maternal genetic score + maternal clinical (intrauterine environment) features:** The maternal genetic score was added to Model 2 to investigate the additional contribution of maternal genetics to variance explained in birthweight.

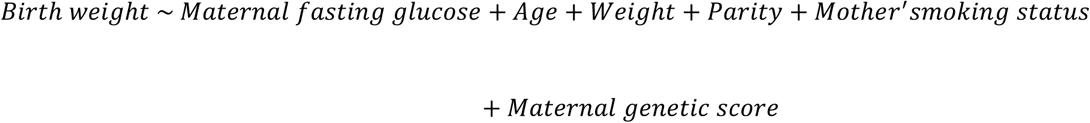

**Model 4: Maternal clinical features + Parental anthropometric traits (genetics) model:** Maternal and paternal height are variables that capture the effects of fetal genetics and are easily measurable; these were added as predictors to Model 2 to create Model 4.

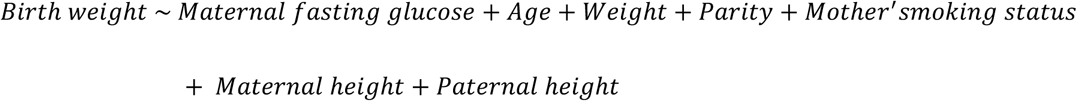

**Model 5: Fetal genetic score + maternal clinical features + parental anthropometric (genetic) traits:** The fetal genetic score was added to Model 4 to further investigate the contributions of the fetal genetic score in addition to parental heights and clinical features.

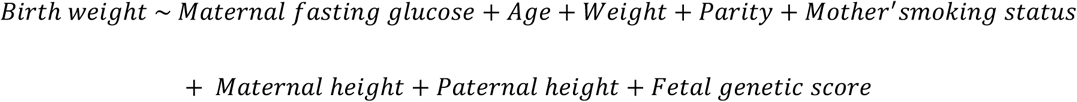

**Model 6: Parental genetic scores + maternal clinical features + parental anthropometric (genetic) traits:** Given that the fetal genetic score for birthweight is not available prior to delivery, we analysed the contribution of the maternal genetic score for offspring birthweight and the paternal genetic score for father’s own birthweight in Model 6 in addition to clinical features and parental heights.

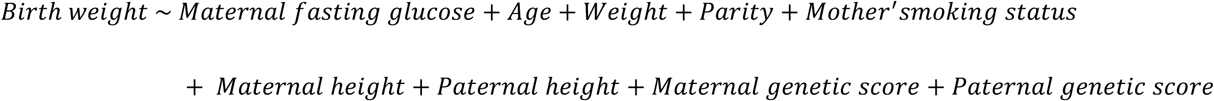

**Model 7: Fetal genetic score + maternal genetic score + maternal clinical features + parental anthropometric (genetic) traits:**

The maternal genetic score was added to Model 5 to further investigate the contributions of the maternal genetic score in addition to parental heights and clinical features.

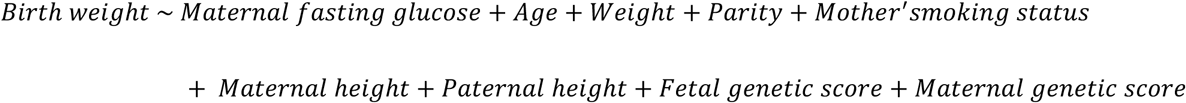

**Additional models: Parents’ own birthweights**

We additionally investigated the contribution of maternal and paternal self-reported birthweights because these may also capture information about fetal genetics. These were available in a smaller sample (n=425 trios).

We used the Adj-R^2^ statistic to assess improvement in model fit based on any added predictors. An F-test was used to compare nested models and check for any improvements in the explanation of variance in birthweight. The R^2^ statistic and its 95% confidence intervals were used to assess the overall explanation of variance in birthweight by the predictors in the model. Confidence intervals were calculated by bootstrapping. Multicollinearity between predictor variables in the models was checked by using the Variance Inflation Factor (VIF).

As a sensitivity analysis to check for any potential impact of poor-quality SNP genotype data, we repeated models containing genetic scores with only those SNPs that had minor allele frequency > 0.1% and imputation quality r^2^ > 0.4. We used the statistical software R (version 3.5.2) to develop the multiple linear regression models and to calculate the F-tests between nested models.

## Results

Descriptive characteristics for the 549 parent-offspring trios are shown in Table 1. There was no strong evidence that individuals excluded from the analysis differed in their basic characteristics from those included (see **Table S2**).

**Table 1:** Key characteristics of study population

| n = 549 trios |  |
| --- | --- |
| Phenotype | Mean or % (SD) |
| Maternal Height (cm) | 165.0 (6.4) |
| Maternal Weight (kg) | 76.3 (12.6) |
| Gestational Duration (weeks) | 40.1 (1.2) |
| Birthweight (g) | 3570 (444) |
| Maternal Age (years) | 30 (5) |
| Maternal smoking status (%Yes) | 14.6 |
| Parity (%1<sup>st</sup> pregnancy) | 44.8 |
| Sex of the baby (% Male) | 52.2 |

The genetic scores all showed strong associations with their respective phenotypes (**Table S3**).

### Maternal and fetal genetic scores contribute additively to offspring birthweight variation

A multivariable linear regression model (Model 1; **Table 2**) showed that maternal and fetal genetic scores have additive contributions to variance in offspring birthweight. On its own, the fetal genetic score explained 2% of variation in adjusted birthweight (R^2^ = 0.020) and the maternal genetic score explained 3% of variance in birthweight (R^2^ = 0.030). For comparison, the variables parity, mother’s smoking status and paternal height each explained 3 % of variation.

**Table 2:** Model 1**-**Results of a multivariable linear regression model testing the association between birthweight (adjusted for sex and gestational age), maternal genetic score and fetal genetic score (n=549 parent-offspring trios). R^2^ = 0.060; Adj-R^2^=0.053

| Variable | Change in birthweight (g) per 1 SD change in independent variable | 95% Confidence Interval | t value | p-value |
| --- | --- | --- | --- | --- |
| Intercept | 3672 | 3530,3813 | 50.9 | <0.001 |
| Maternal genetic score for offspring birthweight (adjusted for fetal effects) | 81 | 45,116 | 4.4 | <0.001 |
| Fetal genetic score for offspring birthweight (adjusted for maternal effects) | 69 | 33,105 | 3.7 | <0.001 |

### Maternal genetic score for birthweight explained additional variance in birthweight when added to easily measurable clinical variables

A multivariable linear regression model (Model 2; **Table 3)** including variables that are readily available in the clinical setting (maternal fasting glucose, maternal age, maternal weight, parity and the mother’s smoking status), showed that each variable contributed to variance explained in birthweight. The total variation in birthweight explained by these maternal characteristics (R^2^ = 0.217) was higher than that explained by genetic scores alone (R^2^ = 0.06; Model 1).

**Table 3:**
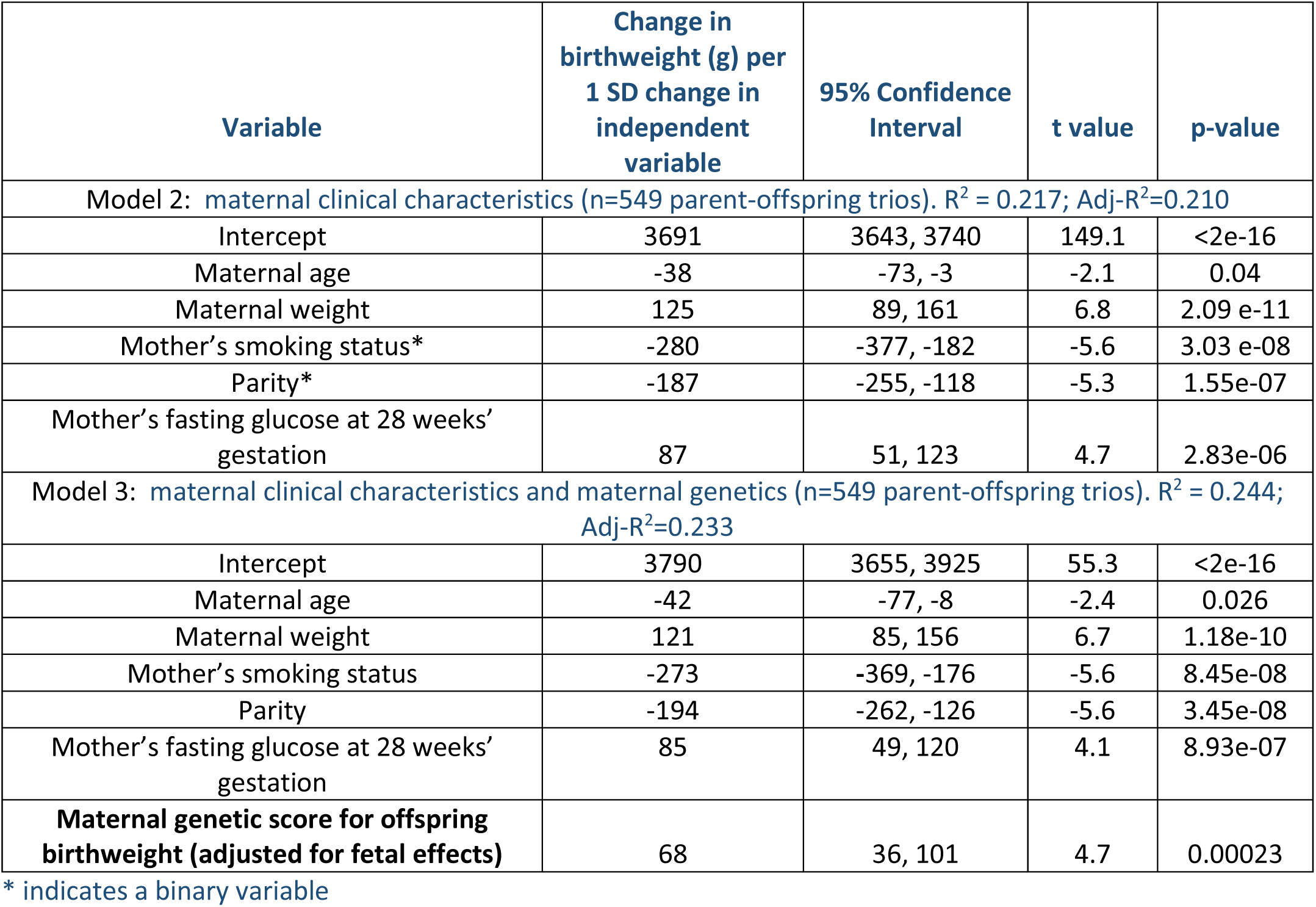
Results of multivariable linear regression models testing the association between birthweight (adjusted for sex and gestational age) and maternal clinical characteristics, with and without the maternal genetic score

The addition of the maternal genetic score for offspring birthweight to Model 2 as a predictor (Model 3; **Table 3**) made little change to the coefficients of the maternal clinical variables, which were very similar to Model 2, but there was an improvement in the Adj-R^2^ statistic when comparing the nested models (Adj-R^2^ =0.233 vs 0.210, p<0.001), indicating that the maternal genetic score captured additional variance in birthweight.

### Maternal and paternal height explained additional variance in birthweight when added to maternal clinical variables

The addition of maternal and paternal height variables, that can capture the effects of fetal genetics, to Model 2 (routinely available clinical features only) showed that the additional variables can further explain variance in birthweight (adjusted for sex and gestational age) (Model 4; **Table 4**) with Adj-R^2^ increasing from 0.210 to 0.248 (p<0.001).

**Table 4:** Results of multivariable linear regression models testing the association between birthweight (adjusted for sex and gestational age), maternal clinical characteristics and parental heights, with and without the fetal genetic score (n=549 parent-offspring trios).

| Variable | Change in birthweight (g) per 1 SD change in independent variable | 95% Confidence Interval | t value | p-value |
| --- | --- | --- | --- | --- |
| Model 4: maternal clinical characteristics and parental heights (n=549 parent-offspring trios). $R^2 = 0.258$; Adj-$R^2=0.248$ | | | | |
| Intercept | 3697 | 3650, 3744 | 152.7 | <2e-16 |
| Maternal age | -49 | -84, -15 | -2.8 | 0.00547 |
| Maternal weight | 101 | 64, 139 | 5.3 | 1.47e-07 |
| Mother's smoking status | -251 | -348, -155 | -5.1 | 3.85e-07 |
| Parity | -210 | -278, -142 | -6.1 | 2.47e-09 |
| Mother's fasting glucose at 28 weeks' gestation | 104 | 68, 140 | 5.7 | 2.03e-08 |
| **Maternal height** | 52 | 16, 87 | 2.9 | 0.00431 |
| **Paternal height** | 69 | 35, 102 | 4.0 | 6.88e-05 |
| Model 5: maternal clinical characteristics (n=549 parent-offspring trios), parental heights, and fetal genetic score. $R^2 = 0.277$; Adj-$R^2=0.264$ | | | | |
| Intercept | 3799 | 3667, 3931 | 56.5 | <2e-16 |
| Maternal age | -50 | -84, -16 | -2.9 | 0.00427 |
| Maternal weight | 97 | 60, 134 | 5.2 | 3.64e-07 |
| Mother's smoking status | -241 | -336, -146 | -5.0 | 8.65e-07 |
| Parity | -216 | -284, -149 | -6.3 | 6.43e-10 |
| Mother's fasting glucose at 28 weeks' gestation | 106 | 71, 142 | 5.9 | 7.42e-09 |
| Maternal height | 49 | 14, 84 | 2.7 | 0.00681 |
| Paternal height | 66 | 33, 99 | 3.9 | 0.000121 |
| **Fetal genetic score for offspring birthweight (adjusted for maternal effects)** | 56 | 23, 88 | 3.4 | 0.000746 |

In a subsample of n=425 available trios, we found that mother’s and father’s own self-reported birthweights explained additional variance in offspring birthweight when added to a model that included parental heights (**Table S4**, Adj-R^2^=0.302 vs 0.258 without parent birthweights, p<0.001).

### Fetal genetic score for birthweight explained additional variance in birthweight when added to easily-measured anthropometric variables that capture fetal genotype

With the addition of the fetal genetic score for offspring birthweight to Model 4 as a predictor (Model 5; **Table 4**), there was little change in the coefficients of the maternal clinical variables, or of the maternal and paternal heights, which were very similar to Model 4, but there was an improvement in the Adj-R^2^ statistic when comparing the nested models (Adj-R^2^ =0.264 vs. 0.248, p<0.001), indicating that the fetal genetic score captured additional variance in birthweight. The fetal genetic score also improved variance explained in the model containing parental birthweights in a subsample of 425 trios (**Table S5**; P=0.09 comparing Adj-R^2^=0.302 for the model with no fetal genetic score with Adj-R^2^= 0.310 for the model with the fetal genetic score).

### Maternal and paternal genetic scores further improved variance explained in birthweight when added to clinical and anthropometric variables

When we added the maternal and paternal genetic scores to Model 4, (Model 6**; Table 5**), both parental genetic scores explained variation in birthweight on top of the basic clinical and anthropometric variables (Adj-R^2^=0.271 vs Adj-R^2^=0.248, p<0.001).

**Table 5:** Model 6-Results of a multivariable linear regression model testing the association between birthweight (adjusted for sex and gestational age), maternal clinical characteristics (n=549 parent-offspring trios), and parental heights and genetic scores. R^2^ = 0.285; Adj-R^2^=0.271

| Variable | Change in birthweight (g) per 1 SD change in independent variable | 95% Confidence Interval | t-value | p-value |
| --- | --- | --- | --- | --- |
| Intercept | 3796 | 3665, 3927 | 56.7 | <2e-16 |
| Maternal age | -54 | -88, -20 | -3.1 | 0.00215 |
| Maternal weight | 97 | 60, 134 | 5.1 | 3.78e-07 |
| Mother's smoking status | -241 | -335, -146 | -5.0 | 8.45e-07 |
| Parity | -211 | --278, -144 | -6.2 | 1.25e-09 |
| Mother's fasting glucose at 28 weeks' gestation | 104 | 69, 140 | 5.8 | 1.50e-08 |
| Maternal height | 44 | 9, 80 | 2.5 | 0.0138 |
| Paternal height | 61 | 28, 95 | 3.6 | 0.000349 |
| **Maternal genetic score for offspring birthweight (adjusted for fetal effects)** | 57 | 25, 90 | 3.5 | 0.000567 |
| **Paternal genetic score for father's own birthweight** | 39 | 6, 71 | 2.3 | 0.0192 |

### Maternal genetic score further improved variance explained in birthweight when added to fetal genetic score

When the maternal genetic score and the fetal genetic scores were added on top of clinical variables (Model 7; **Table 6**), in Model 4 there was additional improvement in explanation of variance in birthweight (Adj-R^2^=0.280 vs Adj-R^2^=0.248, p<0.001).

**Table 6:**
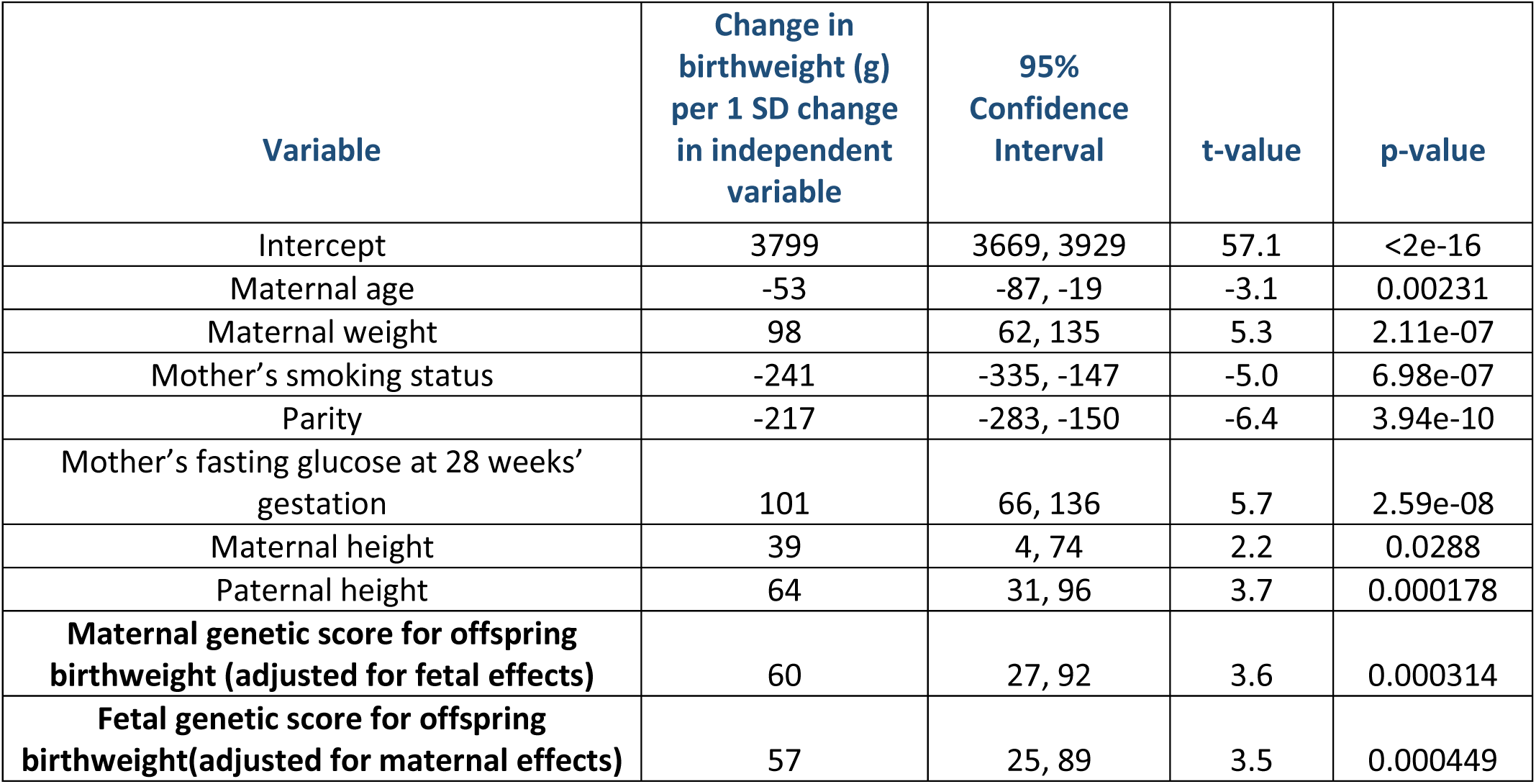
Model 7-Results of a multivariable linear regression model testing the association between birthweight (adjusted for sex and gestational age), maternal clinical characteristics (n=549 parent-offspring trios), parental heights, and maternal and fetal genetic scores. R^2^ = 0.294; Adj-R^2^=0.280

| Variable | Change in birthweight (g) per 1 SD change in independent variable | 95% Confidence Interval | t-value | p-value |
| --- | --- | --- | --- | --- |
| Intercept | 3799 | 3669, 3929 | 57.1 | <2e-16 |
| Maternal age | -53 | -87, -19 | -3.1 | 0.00231 |
| Maternal weight | 98 | 62, 135 | 5.3 | 2.11e-07 |
| Mother's smoking status | -241 | -335, -147 | -5.0 | 6.98e-07 |
| Parity | -217 | -283, -150 | -6.4 | 3.94e-10 |
| Mother's fasting glucose at 28 weeks' gestation | 101 | 66, 136 | 5.7 | 2.59e-08 |
| Maternal height | 39 | 4, 74 | 2.2 | 0.0288 |
| Paternal height | 64 | 31, 96 | 3.7 | 0.000178 |
| **Maternal genetic score for offspring birthweight (adjusted for fetal effects)** | 60 | 27, 92 | 3.6 | 0.000314 |
| **Fetal genetic score for offspring birthweight(adjusted for maternal effects)** | 57 | 25, 89 | 3.5 | 0.000449 |

A summary of the R^2^ values across all the main models is shown in **Figure 2**. This indicates the improvement in R^2^ with added successive variables.

**Fig 2:**
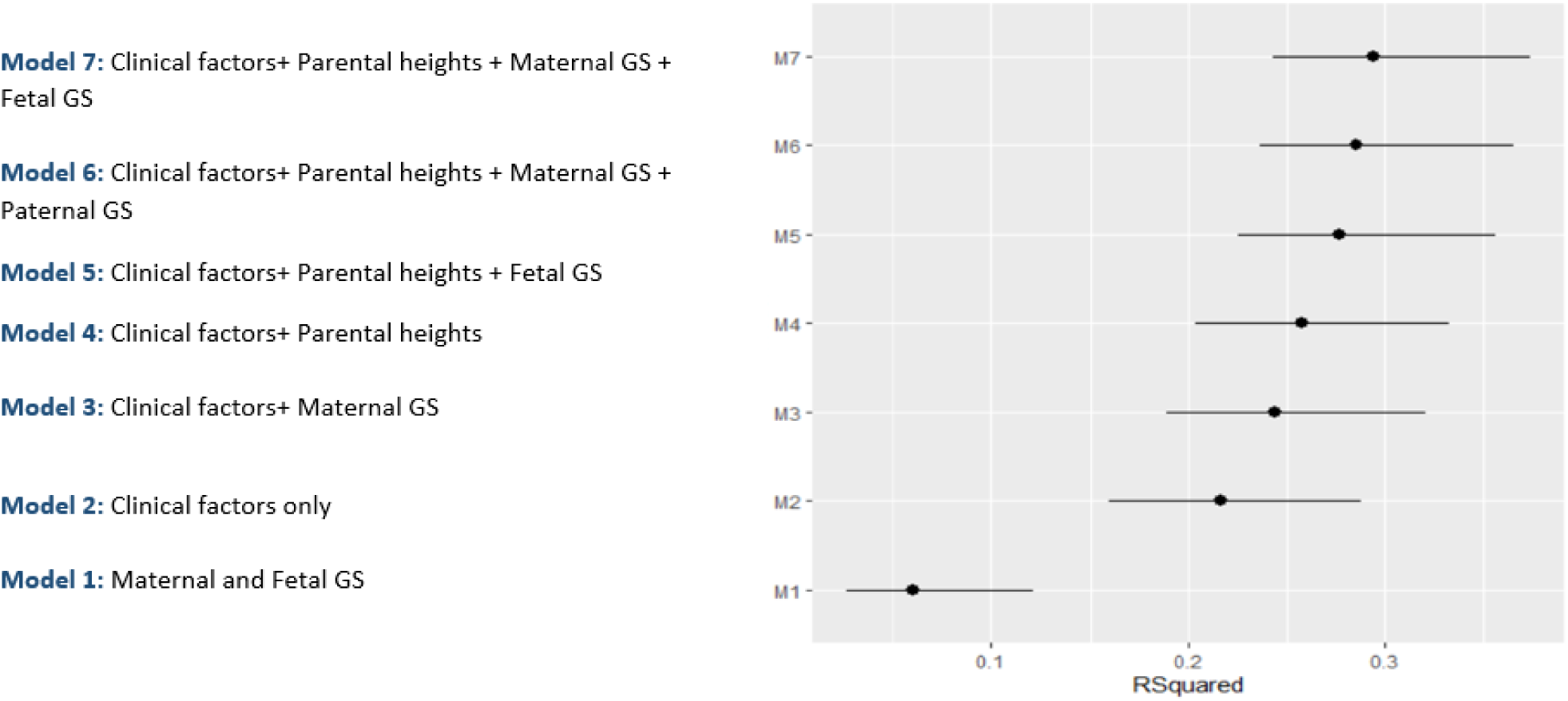
Plot showing R-squared values for each model with 95% confidence intervals.

There was a negligible difference between the models that contained genetic scores with only those SNPs that had minor allele frequency > 0.1% and imputation quality r^2^ > 0.4 and the models that contained genetic scores with SNPs having minor allele frequency > 0.001 and imputation quality > 0.4.

## Discussion

We have shown that maternal, paternal and fetal genetic scores contribute to variation in sex- and gestational age-adjusted birthweight, in addition to variables easily obtained in a clinical setting. We also showed that maternal and paternal heights, which are easily measured and capture some of the genetic contribution to fetal growth, explain variance in birthweight independently of routinely measured maternal clinical variables. However, the maternal and fetal (or paternal) genetic scores made additional, independent contributions to birthweight variance. GWAS have established that fetal and maternal genetic variants are associated with birthweight^12^, but as many of the underlying causal genes are likely associated with clinical or anthropometric traits, such as height, weight, and glucose, it has been important to quantify the added value.

Maternal and fetal genetics are known to be important determinants of fetal growth but the contribution of genetic scores to variance explained in birthweight has not been investigated previously using multivariable regression models containing other clinical and parental anthropometric characteristics. We showed, consistent with other epidemiological studies^18, 19^, that clinical variables, both routinely measured (glucose, weight, smoking), but also parental height, can explain approximately 26% variation in birthweight that has already been adjusted for sex and gestational age. The addition of the fetal genetic score to the models explained a further 2% of variation in birthweight. For comparison, the variables parity, mother’s smoking status and paternal height each explained 3 % of variation individually, in sex-and gestational age-adjusted birthweight. The precise mechanisms through which most of the genetic variants in the fetal score influence growth are not known, but evidence to date suggests they are likely to capture variation in growth factors such as fetal insulin, as well as variation in placental growth and function^12^.

Fetal genetic scores are not available before birth, so they are not informative for predicting birthweight at present. However, we showed that maternal and paternal genetic scores can also explain variation in birthweight. The parental genetic effects are mediated both through direct effects of genes inherited by the fetus and indirect maternal genetic effects on the intra-uterine environment. Some of these effects will have been captured by clinical features. Previous research has shown that associations between maternal height and offspring birthweight is predominantly defined by fetal genetics^20^. Paternal height has also been shown to influence offspring birthweight through fetal genetics^14^. We have shown that the parental heights explain further variation in birthweight and that parental genetic scores for birthweight are contributing to variation in birthweight independently of parental heights. The independent and additive associations of the parental genetic scores with birthweight show that these scores are offering additional predictive value. The fetal genetic score also added information on top of self-reported parent birthweights.

It was unexpected that the R^2^ value for the maternal GS was larger than that of the fetal GS because previous work ^12^ has shown that fetal genetic variants explain more birthweight variation than maternal genetic variants. However, further investigation showed that the R^2^ values for maternal and fetal genetic scores were not precise enough in this relatively small sample to be able to infer confidently whether one was bigger than the other (as reflected in the 95% confidence intervals), and point estimate values of R^2^ fluctuated so that the fetal estimate appeared larger than the maternal estimate when the models were re-run in wider samples that did not require all family members to be genotyped (see **Table S6**).

This study has benefited from the use of a well-phenotyped and genotyped sample of parents and children. However, there are some limitations. Firstly, in the EFSOCH dataset, some clinical features known to contribute to variance explained in birthweight in other studies (e.g. blood pressure ^4^) were not available, so studies in additional samples would be needed to enable assessment of the contribution of genetic scores in relation to those variables. In addition, although we aimed to assess the contribution of parents’ own birthweights as anthropometric variables in addition to parental heights, the parental birthweights were self-reported and were not available in the full sample (they were available in only 425 complete trios). However, when we created models using the dataset containing 425 trios (**Tables S4-5**), the coefficients of the explanatory variables were similar to those in the models created with the larger dataset of 549 observations, so the limited availability of self-reported birthweights did not impact materially on the results.

Another limitation of this study is that we conducted the analyses in a UK-based, northern European-ancestry population and it is likely that the associations between birthweight and both genetics and parental clinical features will differ in samples of other ancestries and in other settings. Further studies will be necessary to investigate the contribution of genetic scores and other variables to birthweight in other populations.

Since the EFSOCH study was part of the maternal GWAS study that identified SNPs associated with birthweight^12^, there is a small risk of overfitting in our models. However, we expect the risk of this to be minimal because EFSOCH only made up 0.4% of the maternal GWAS meta-analysis sample and was not included in the fetal GWAS.

We have shown that maternal and fetal genetic scores explain variation in birthweight in healthy pregnancies, in addition to clinical and anthropometric variables that are routinely or easily collected. While the individual contribution of each genetic score is not large (e.g. 2% for fetal genetic score), it is comparable to the individual contributions of variables such as parity or maternal smoking status. This raises the possibility that genetic scores might be useful alongside clinical characteristics in prediction models, for example, those aiming to predict risk of LGA in pregnancies affected by gestational diabetes. Further work is needed to determine whether genetic information could improve a full clinical prediction model over and above what is currently done routinely in clinical practice.

## Data Availability

Requests for access to the original EFSOCH dataset should be made in writing in the first instance to the EFSOCH data team via the Exeter Clinical Research Facility.
GWAS summary statistics for birthweight that were used to generate the genetic scores are publicly available can be downloaded from http://egg-consortium.org/

http://egg-consortium.org/

## Ethics approval

Ethical approval for the Exeter Family of Childhood Health was given by the North and East Devon (UK) Local Research Ethics Committee (approval number 1104), and informed consent was obtained from the parents of the new-borns.

## Data Availability

Requests for access to the original EFSOCH dataset should be made in writing in the first instance to the EFSOCH data team via the Exeter Clinical Research Facility.

GWAS summary statistics for birthweight that were used to generate the genetic scores are publicly available can be downloaded from http://egg-consortium.org/

## Acknowledgements

We would like to thank all the families who contributed to The Exeter Family Study of Childhood Health.

## Supplementary tables

**Table S2:**
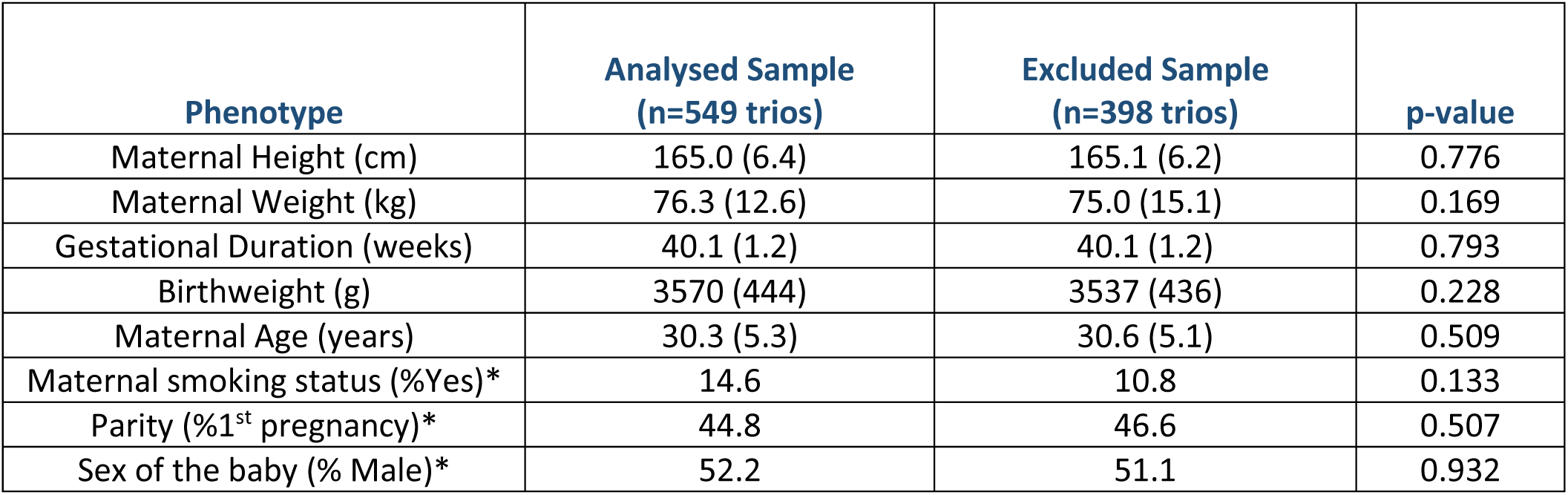
Key characteristics of excluded participants and results of t-tests and chi-square tests for comparison between the analysed sample and the excluded sample. * indicates the p-value is for a Chi Square test

**Table S3:** Associations of maternal, fetal and paternal genetic scores with birthweight

| Outcome Variable | Genetic score | Change in trait per 1 SD higher genetic score | 95% CI | Pearson's Correlation coefficient, r | p-value | n |
| --- | --- | --- | --- | --- | --- | --- |
| Offspring birthweight adjusted for sex and gestational age | Maternal genetic score for offspring birthweight (g), with weights adjusted for fetal genotype effects | 80.4g | 43.7,117.1 | 0.18 | 2.03e-05 | 549 |
| Offspring birthweight adjusted for sex and gestational age | Fetal genetic score for birthweight (g), with weights adjusted for maternal genotype effects | 68.0 g | 31.3,104.9 | 0.15 | 0.000303 | 549 |
| Father's own self-reported birthweight | Paternal genetic score for father's own birthweight (generated with unadjusted weights) | 59.2g | 27.8, 90.7 | 0.17 | 0.00024 | 504 |

**Table S4:** Results of a multivariable linear regression model testing the association between birthweight (adjusted for sex and gestational age), maternal clinical characteristics (n=425 parent-offspring trios), and additional parental anthropometric features that capture fetal genetics. R^2^=0.317; Adj-R^2^=0.302

| Variable | Change in birthweight (g) per 1 SD change in X variable | 95% Confidence Interval | t value | p-value |
| --- | --- | --- | --- | --- |
| Intercept | 3727 | 3654, 3778 | 142.9 | <0.001 |
| Maternal age | -44 | -81, -7 | -2.3 | 0.02 |
| Maternal weight | 82 | 41, 122 | 4.0 | <0.001 |
| Mother's smoking status | -237 | -350, -123 | -4.1 | <0.001 |
| Parity | -219 | -291, -147 | -6.0 | <0.001 |
| Mother's fasting glucose at 28 weeks' gestation | 105 | 67, 144 | 5.3 | <0.001 |
| **Mother's own birthweight** | 103 | 66, 140 | 5.5 | <0.001 |
| **Father's own birthweight** | 47 | 10, 83 | 2.5 | 0.01 |
| Maternal height | 23 | -16, 62 | 1.2 | 0.25 |
| Paternal height | 60 | 23, 98 | 3.2 | 0.002 |

**Table S5:** Results of a multivariable linear regression model testing the association between birthweight (adjusted for sex and gestational age), maternal clinical characteristics (n=425 parent-offspring trios), additional features that capture fetal genetics, and fetal genetic score. R^2^ = 0.329; Adj-R^2^=0.310, (p=0.06 when compared to Model in table S4)

| Variable | Change in birthweight (g) per 1 SD change in X variable | 95% Confidence Interval | t value | p-value |
| --- | --- | --- | --- | --- |
| Intercept | 3870 | 3724, 4016 | 51.9 | <0.001 |
| Maternal age | -43 | -80, -6 | -2.3 | 0.02 |
| Maternal weight | 81 | 41, 122 | 4.0 | <0.001 |
| Mother's smoking status | -230 | -343, -117 | -4.0 | <0.001 |
| Parity | -226 | -298, -154 | -6.1 | <0.001 |
| Mother's fasting glucose at 28 weeks' gestation | 105 | 66, 143 | 5.3 | <0.001 |
| Mother's own birthweight | 94 | 57, 132 | 4.9 | <0.001 |
| Father's own birthweight | 46 | 9, 82 | 2.5 | 0.01 |
| Maternal height | 23 | -16, 61 | 1.1 | 0.25 |
| Paternal height | 60 | 23, 97 | 3.2 | 0.002 |
| **Fetal genetic score for offspring birthweight (adjusted for maternal effects)** | 32 | -4, 67 | 1.8 | 0.080 |

**Table S6:** Summary of models describing the contribution of the genetic scores to variation in offspring birthweight in different sample sizes.

| Included sample (of those with birthweight and complete phenotype data available) | Total N | Description of model | Change in BW in g per 1 SD higher X variable (95%CI) | R<sup>2</sup> (95%CI) |
| --- | --- | --- | --- | --- |
| Max sample with maternal genetic score | 820 | Outcome: birthweight (adjusted for sex and gestational age)<br>X variable: maternal genetic score | 66 (32-100) | 0.02432<br>(0.00513, 0.0502) |
| Max sample with fetal genetic score | 646 | Outcome: birthweight (adjusted for sex and gestational age)<br>X variable: fetal genetic score | 84 (50-117) | 0.03634<br>(0.0131, 0.0710) |
| Max sample with maternal and fetal genetic scores | 595 | Outcome: birthweight (adjusted for sex and gestational age)<br>X variable: maternal genetic score | 73 (38-108) | 0.02935<br>(0.00733, 0.0595) |
| Max sample with maternal and fetal genetic scores | 595 | Outcome: birthweight (adjusted for sex and gestational age)<br>X variable: fetal genetic score | 78(43-113) | 0.03307<br>(0.00941, 0.0654) |
| Max sample with maternal, fetal and paternal genetic scores | 549 | Outcome: birthweight (adjusted for sex and gestational age)<br>X variable: maternal genetic score | 80<br>(44-117) | 0.036<br>(0.00966, 0.0697) |
| Max sample with maternal, fetal and paternal genetic scores | 549 | Outcome: birthweight (adjusted for sex and gestational age)<br>X variable: fetal genetic score | 68<br>(31-105) | 0.0269<br>(0.00497, 0.0557) |

